# A priority index-based computational medicine framework (PimRNA) for prioritising personalised mRNA cancer vaccines

**DOI:** 10.64898/2026.05.26.26354114

**Authors:** Tingting Tan, Hai Fang

## Abstract

**Background:** The development of personalised mRNA cancer vaccines holds considerable promise for oncology, yet a significant translational gap persists between neoantigen identification and the selection of therapeutically impactful targets. Current approaches predominantly prioritise human leukocyte antigen (HLA) binding affinity and immunogenicity, often overlooking the systems-level biological context of the target. This can inadvertently favour immunogenic but biologically peripheral peptides that exert limited influence on tumour signalling networks, thereby constraining vaccine efficacy. Furthermore, mRNA therapeutics must satisfy additional design requirements, including favourable codon usage and favourable secondary-structure stability, which directly affect in vivo translation and half-life. A unified computational framework that integrates neoantigen discovery with network biology is therefore critically needed.

**Results:** Here, we present PimRNA, a Priority index (Pi)-centric computational medicine framework that bridges this gap by unifying neoantigen identification, mRNA sequence optimisation, and gene interaction network analysis. First, high-confidence tumour-specific HLA class I and II neoantigenic peptides are identified from paired tumour-normal genomic and tumour transcriptomic data using NeoDisc. Second, the coding sequences of these peptides are optimised for stability and translational efficiency with LinearDesign, yielding a core set of neoantigen-encoding mRNAs. Third, a random walk with restart algorithm is applied to a knowledgebase of gene interactions to identify peripheral genes exhibiting significant network connectivity to core genes, generating a gene-predictor matrix in which each gene is assigned an affinity score reflecting its network proximity to immunogenic neoantigens. These scores are consolidated into a single, unified priority rating (0-5) for each gene, followed by subnetwork analysis that reveals therapeutically relevant gene modules. Application of PimRNA to breast cancer and melanoma datasets demonstrates that it successfully selects high-confidence immunogenic neoantigen candidates embedded within biologically meaningful tumour-specific networks.

**Conclusion:** PimRNA provides a systems biology foundation for mRNA vaccine design, moving beyond isolated immunogenicity to prioritise targets that are both highly presented and central to tumour-relevant biological networks. This framework offers a generalisable strategy for the rational discovery and prioritisation of mRNA therapeutics, significantly advancing the field of computational medicine towards personalised cancer vaccines.

**Key Points:**

- PimRNA integrates neoantigen discovery, mRNA sequence optimisation, and gene interaction network analysis into a single computational medicine framework.
- A random walk with restart algorithm identifies peripheral genes with strong network connectivity to core genes defined by optimised neoantigen-encoding mRNAs, and Fisher’s combined meta-analysis consolidates network-based affinity scores into a unified priority rating (0–5) for each gene, enabling rational target prioritisation.
- Application to breast cancer and melanoma demonstrates that PimRNA selects immunogenic neoantigens that are also central to tumour-relevant signalling networks, moving beyond isolated binding predictions.
- The framework provides a generalisable, systems biology-driven strategy for the design and prioritisation of mRNA cancer vaccines.

## Introduction

The advent of mRNA therapeutics has fundamentally reshaped precision medicine, with personalised cancer vaccines representing one of the most transformative applications to emerge from this technological revolution^1,2^. By leveraging *in vitro*-synthesised mRNA to encode patient-specific tumour neoantigens, these vaccines induce tailored T-cell responses that selectively target malignant cells while sparing healthy cells^3^. The clinical promise of this approach has been substantiated by recent trials, including the personalised mRNA vaccine autogene cevumeran in pancreatic ductal adenocarcinoma, where vaccine-induced CD8□T-cell responses correlated with significantly prolonged recurrence-free survival^3,4^. These landmark studies underscore the potential of neoantigen-directed mRNA vaccines to improve outcomes in genomically complex malignancies.

The platform’s clinical relevance is particularly compelling in breast cancer, especially triple-negative breast cancer (TNBC), whose pronounced genomic instability and immune-inflamed tumour microenvironment render it an especially attractive target for mRNA-based immunotherapy. A phase I study of an individualised neoantigen mRNA vaccine delivered via lipid nanoparticles in adjuvant TNBC demonstrated durable T-cell immunity and prolonged relapse-free survival, with most patients remaining disease-free at extended follow-up^5^. These vaccines are constructed using computational pipelines and artificial intelligence to identify patient-specific neoantigens, and further refinements in mRNA sequence design and lipid nanoparticle formulation have improved stability, translational efficiency and tumour targeting^6^. Beyond vaccination, mRNA technology now enables the in vivo generation of chimeric antigen receptor T cells directed against TROP2, producing clinical tumour regression in breast cancer without the need for ex vivo cell manufacturing^7^. Preclinical studies have additionally shown that mRNA-mediated restoration of tumour-suppressor proteins—including p53, PTEN and PTPN14—can reverse malignant phenotypes and suppress disease progression in TNBC models^8,9^. Collectively, these advances position mRNA therapeutics as a versatile, personalised modality capable of complementing chemoimmunotherapy and antibody–drug conjugates, particularly in hard-to-treat subtypes such as TNBC^10^.

Central to the success of personalised mRNA cancer vaccines is the accurate identification and prioritisation of tumour-specific neoantigens—mutated peptides that arise from somatic alterations, are presented by human leukocyte antigen (HLA) molecules, and are recognised by the adaptive immune system. Contemporary computational pipelines, including NeoDisc^11^ and pVACtools^12^, have streamlined neoantigen discovery by integrating whole-genome or whole-exome sequencing, transcriptomic confirmation of mutant gene expression, HLA typing, peptide–MHC binding affinity prediction, and immunogenicity assessment into end-to-end workflows. These tools have markedly accelerated the transition from tumour biopsy to vaccine candidate identification, reducing what was once a laborious low-throughput process to a matter of days. Despite these advances, however, a critical translational gap persists: current approaches overwhelmingly prioritise neoantigens based on predicted HLA binding affinity and T-cell recognition potential, often at the expense of systems-level biological context.

Compounding this limitation, existing pipelines are almost exclusively designed to detect somatic mutations within canonical protein-coding genes, thereby overlooking an entirely distinct reservoir of tumour antigens derived from non-canonical open reading frames (ncORFs) in the so-called ‘dark proteome’. A transformative new class of antigens—termed peptideins—has recently been described by the TransCODE Consortium, originating from ncORFs within genomic regions long regarded as non-coding, including untranslated regions, long non-coding RNAs and overlapping ORFs^13^. Although these translation products have been experimentally confirmed, their full functional characterisation as classical proteins remains incomplete. Critically, peptideins are actively processed and presented via HLA class I molecules, conferring genuine immunological visibility and establishing them as cryptic neoantigens. This discovery substantially expands the targetable immunopeptidome well beyond canonical protein-coding genes. The enrichment of peptideins is particularly pronounced in cancers, including paediatric tumours and low-mutation-burden malignancies, where conventional neoantigen discovery is constrained, indicating that a large reservoir of tumour-specific immune targets has been overlooked. Peptideins therefore revise foundational assumptions in neoantigen screening, cancer immunotherapy and genomic variant interpretation, as alterations in non-coding regions previously dismissed as silent may now be understood to exert pathogenic effects through peptidein expression.

Even when neoantigens are accurately identified, the prevailing immunogenicity-centric paradigm carries an inherent limitation. By focusing narrowly on immunogenic peptides in isolation, existing pipelines may select targets that, whilst capable of eliciting a measurable T-cell response, exert limited influence on the signalling networks that sustain tumour growth and immune evasion. A neoantigen derived from a biologically peripheral gene—one that is not central to cancer-relevant pathways— may fail to disrupt the tumour’s functional architecture, thereby constraining vaccine efficacy. Conversely, targeting genes that occupy hub positions within tumour-specific interaction networks could amplify therapeutic impact by destabilising multiple downstream oncogenic processes. Crucially, such network-central nodes are often subject to higher evolutionary constraint and represent tumour-essential programmes whose loss incurs a substantial fitness cost, rendering them less susceptible to immune escape through antigen downregulation.

This biological logic not only supports prioritising neoantigens from hub genes but also opens the door to a complementary strategy: the inclusion of biologically important yet previously under-ranked candidates that occupy key network positions. These neoantigens may be co-delivered with highly immunogenic neoantigens in a multivalent mRNA vaccine. By targeting network vulnerabilities that the tumour cannot readily discard, such a strategy provides an additional safeguard against immune evasion, reinforcing the durability of vaccine-induced responses. The concept of “network vulnerability”—whereby disrupting central nodes yields disproportionate therapeutic effects—has gained traction in oncology, yet it has not been systematically integrated into the mRNA neoantigen prioritisation process, and the potential of network-informed neoantigens remains largely untapped.

Beyond immunological considerations, the design of mRNA therapeutics also requires attention to sequence-level properties that directly influence clinical translatability. The open reading frame must be codon-optimised to balance translational efficiency with structural stability, avoiding excessive GC content that promotes deleterious secondary structures whilst maintaining sufficient thermostability to resist nuclease degradation. Untranslated regions require engineering to enhance mRNA half-life and ribosome recruitment, and the overall sequence must minimise innate immune activation through strategic nucleotide modifications or removal of double-stranded RNA contaminants. Tools such as LinearDesign^14^ have demonstrated that simultaneous optimisation of minimum free energy and codon adaptation index can markedly increase protein expression and in vivo antibody titres; however, such sequence optimisation is typically applied to candidate antigens only after they have been selected through immunology-centric pipelines. This sequential workflow—neoantigen identification followed by sequence optimisation—does not currently incorporate network biological information at the selection stage, representing an opportunity for methodological integration.

Here, we present PimRNA, a Priority index (Pi)-centric computational medicine framework designed to bridge the gap between neoantigen discovery and systems-informed target prioritisation for personalised mRNA cancer vaccines^15–17^. Our approach unifies three previously disparate components: high-confidence neoantigen identification from paired tumour-normal genomic and tumour transcriptomic data; mRNA sequence optimisation for stability and translational efficiency; and gene interaction network analysis to identify targets that are both immunogenic and centrally positioned within tumour-relevant biological networks. Specifically, we employ NeoDisc to identify HLA class I and II neoantigenic peptides, whose mRNA sequences are subsequently optimised using LinearDesign to generate a core set of neoantigen-encoding genes. A random walk with restart algorithm is then applied to a knowledgebase of gene interactions to identify peripheral genes exhibiting significant network connectivity to this core set. Fisher’s combined meta-analysis consolidates network-based affinity scores into a unified priority rating scaled from 0 to 5 for each gene, enabling rational subnetwork analysis that reveals therapeutically relevant gene modules comprising biologically important yet under-ranked candidates. As illustrated through its application in breast cancer and melanoma datasets, PimRNA successfully selects high-confidence immunogenic candidates embedded within biologically meaningful tumour-specific networks, moving beyond isolated binding predictions to embrace a systems biology foundation for mRNA vaccine design.

## Methods

### Neoantigen identification and mRNA sequence optimisation

PimRNA builds upon the Pi concept to unify neoantigen discovery with network biology. Paired tumour-normal genomic and tumour transcriptomic data^18,19^ from a breast cancer patient (#4136) and a melanoma patient (#2369) were first processed with NeoDisc to identify tumour-specific HLA class I and class II neoantigenic peptides. Peptides with filter annotations other than “Not Selected”, including “Top ranking”, “Driver” and “Not predictable with ML”, were retained for downstream analysis. Their corresponding mRNA coding sequences were optimised for both stability and translational efficiency using LinearDesign, which jointly maximises the codon adaptation index (CAI) and minimises the minimum free energy (MFE) of the folded mRNA. Each optimised sequence was assigned a score defined as CAI × |MFE|. This weighting strategy embeds a manufacturability constraint at the earliest stage of target selection, ensuring that even neoantigens situated at network hubs are penalised if their coding sequences are predicted to have poor translational efficiency (low CAI) or to adopt highly unstable secondary structures (low |MFE|). In this way, the final priority rating reflects both biological importance and the practical feasibility of achieving robust in vivo expression. The corresponding genes of candidate peptides were designated as core genes, stratified into HLA-I core genes and HLA-II core genes. When multiple peptides mapped to the same gene, the peptide with the highest weight was retained as the representative candidate. These scored core genes served as seeds for all subsequent network analyses.

### Network-based peripheral gene identification and predictor matrix construction

A comprehensive gene interaction knowledgebase, termed NetKB, was assembled by merging high-quality functional interactions from STRING^20^ (restricted to the evidence codes “experiments” and “databases”) with pathway-derived gene interactions from KEGG^21^. For each HLA class separately, the scored core genes were used as restarting seeds in a random walk with restart (RWR) algorithm operating on NetKB. The RWR assigned an affinity score on a continuous scale from 0 to 1 to every gene in the network, reflecting its proximity to the core gene set. This procedure generated two genome-wide predictors—the HLA-I predictor and the HLA-II predictor—each comprising both core and peripheral genes together with their affinity scores. The results were organised into a gene-predictor matrix of approximately 16,000 rows (genes) and two columns (the HLA-I and HLA-II predictors).

### Priority rating calculation

Within each predictor, the affinity scores were transformed into P-like values using the empirical cumulative distribution function (eCDF). For a given gene, the two resulting P-like values (one from the HLA-I predictor and one from the HLA-II predictor) were combined by Fisher’s combined method for meta-analysis. Although the HLA-I and HLA-II affinity scores for the same gene may be correlated through shared dependence on transcript abundance, the antigen-processing and recognition pathways they represent are mechanistically independent; Fisher’s method is therefore appropriate for aggregating these two sources of evidence. The combined statistic was then linearly rescaled to yield a priority rating on a continuous scale from 0 to 5, with higher values indicating stronger concordant evidence from both neoantigen presentation and network connectivity.

### Pathway-level subnetwork analysis

To further prioritise targets situated at the intersection of key signalling pathways, a subnetwork of interconnected high-priority genes was extracted from NetKB by focusing on those genes with elevated priority ratings. Within this subnetwork, pathway crosstalk of potential therapeutic relevance was identified by solving a prize-collecting Steiner tree problem heuristically, thereby revealing minimal connected subgraphs enriched in high-priority nodes. The statistical significance of the observed crosstalk was assessed with a degree-preserving node permutation test (100 iterations), yielding an empirical P-value. The final subnetwork was visualised using the stress majorisation algorithm to facilitate biological interpretation and the rational selection of mRNA therapeutic candidates.

### Code availability

Source codes used in the study are accessible following detailed step-by-step instructions at http://www.genetictargets.com/PIMRNA/index.html to reproduce all results, including the figures presented in the paper. Packaged codes have been deposited into a GitHub repository at https://github.com/hfang-bristol/PIMRNA. Our commitment to transparency in code and data sharing promotes reproducible research, allowing the broader clinical and research community to benefit fully from our work without any barriers.

## Results

### PimRNA framework identifies tumour-specific neoantigens with favourable immunogenicity and network-level importance

Current neoantigen discovery pipelines primarily prioritise candidate peptides according to predicted HLA presentation and immunogenicity, yet often lack systems-level evaluation of the biological relevance of the corresponding genes. To address this limitation, we developed PimRNA, an integrated framework that combines neoantigen discovery, mRNA sequence optimisation, and network-based prioritisation to identify candidate neoantigens with both immunological and functional significance (**Figure 1**). Briefly, PimRNA first identifies tumour-specific HLA-I and HLA-II peptides using NeoDisc, followed by mRNA sequence optimisation with LinearDesign and subsequent systems-level prioritisation with Pi.

**Figure 1.**
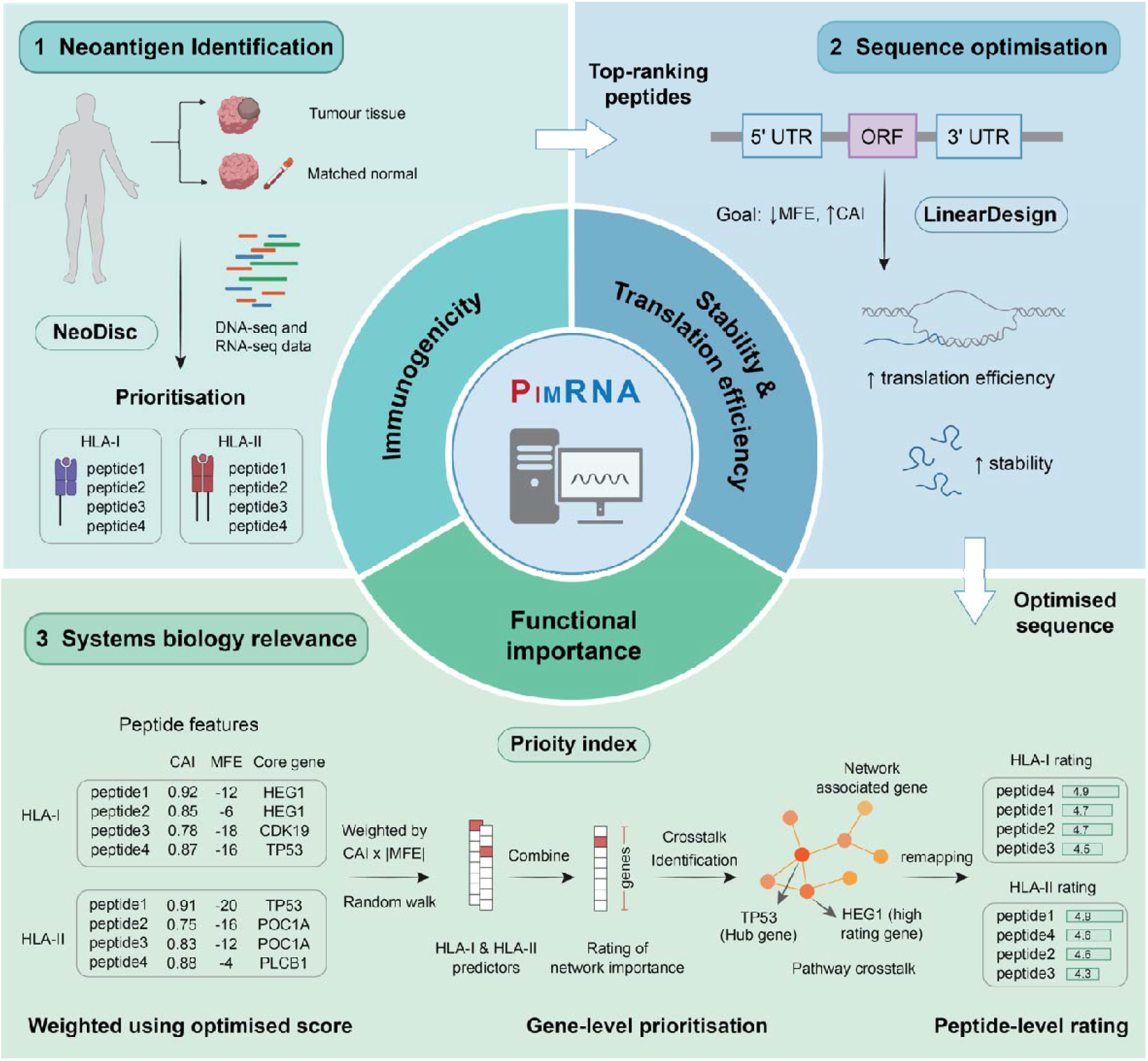
The PimRNA framework for prioritising neoantigen-based mRNA vaccine targets. The schematic outlines a three-stage pipeline. First, neoantigen identification uses NeoDisc on paired tumour-normal genomic and transcriptomic data to find tumour-specific HLA class I and II neoantigenic peptides. Second, mRNA sequence optimisation applies LinearDesign to the coding sequences of the top-ranked peptides, jointly maximising the codon adaptation index and minimising minimum free energy to enhance translational efficiency and stability. Third, systems biology analysis assesses the functional importance of the encoding genes by constructing a core gene set from the optimised sequences and propagating it through a gene interaction knowledgebase using a random walk with restart to assign network-based affinity scores. These scores are combined into a priority rating for gene-level prioritisation. Subsequent crosstalk analysis reveals therapeutically relevant subnetworks comprising high-priority and hub genes, which are then mapped back to constituent peptides to generate the final prioritised list of candidate antigens for multivalent mRNA vaccines.

To evaluate the PimRNA framework, paired tumour-normal genomic and tumour transcriptomic sequencing data from a melanoma patient (#2369) and a breast cancer patient (#4136) were analysed. In the first step, NeoDisc was used to perform somatic mutation calling, HLA typing, peptide presentation prediction, and immunogenicity modelling to generate candidate HLA-I and HLA-II neoantigens (**Figure 2a**). In the melanoma sample, HLA-I peptides constituted the majority of predicted neoantigens, substantially exceeding the number of HLA-II candidates (**Figure 2b**). Predicted mutant peptide binding ranks were generally low for both HLA classes, with many mutant peptides showing improved predicted HLA binding relative to their wild-type counterparts (**Figure 2c,d**), supporting favourable peptide–HLA binding affinity and tumour specificity. Several genes generated multiple independent neoantigen candidates, including *BCR, DDB1, BLTP2, RNPEP, CHD3*, and *BRAF* (**Figure 2e**). Notably, *BRAF* is one of the most extensively characterised oncogenic drivers in melanoma and is strongly linked to MAPK pathway activation and tumour progression^22^. In addition, *DDB1* participates in DNA damage repair and ubiquitin-mediated protein degradation and has been implicated in UV-induced nucleotide excision repair and melanoma-associated genomic instability^23^. These results suggest that NeoDisc captured neoantigen candidates associated with diverse tumour-related biological processes, providing an initial candidate landscape for subsequent network-based prioritisation analyses.

**Figure 2.**
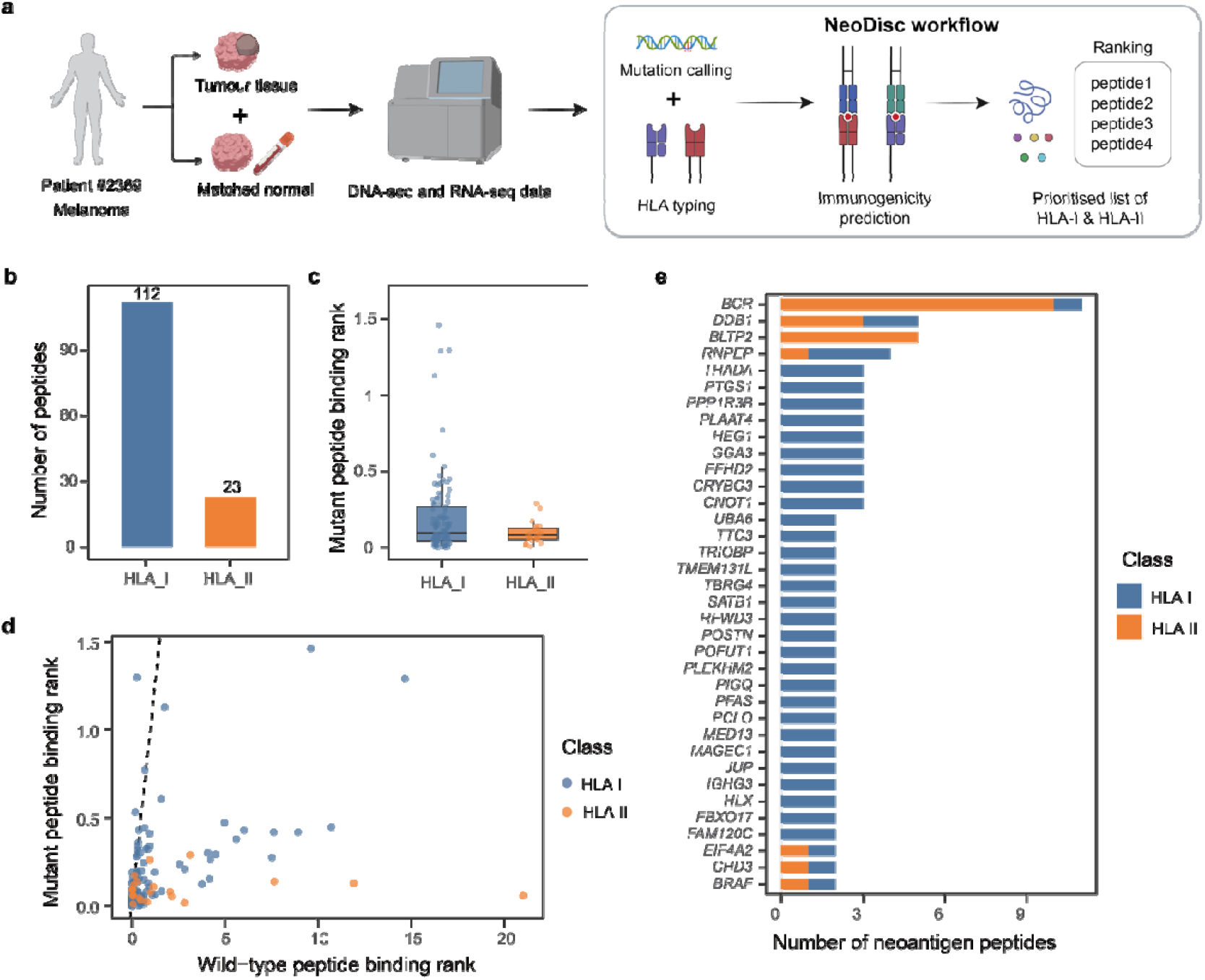
Identification and prioritisation of candidate neoantigens using the NeoDisc pipeline. Panel (a) provides an overview of the NeoDisc workflow, which integrates somatic mutation calling and HLA typing from paired tumour-normal DNA and tumour RNA sequencing data, followed by evaluation of candidate mutant peptides for their HLA-I and HLA-II presentation potential and immunogenicity. Panel (b) displays the total number of predicted HLA-I and HLA-II peptides identified by NeoDisc. Panel (c) illustrates the distribution of mutant peptide binding ranks, where lower values indicate stronger predicted binding affinity. Panel (d) compares mutant peptide binding ranks with corresponding wild-type peptide binding ranks, with the dashed diagonal line representing equal binding affinity. Panel (e) presents the top neoantigen-associated genes ranked by the number of predicted HLA-I and HLA-II neoantigen peptides, with bar colours indicating the peptide class.

### Integrating translational feasibility into neoantigen prioritisation

The translational feasibility of candidate mRNA sequences is rarely considered during early-stage neoantigen target selection. To address this limitation, candidate peptide coding sequences were optimised using LinearDesign (**Figure 3a**), which jointly maximises the codon adaptation index (CAI) and minimises mRNA folding free energy (MFE), thereby improving predicted translational efficiency and transcript stability. The resulting optimisation landscape revealed substantial heterogeneity across neoantigen candidates (**Figure 3b**). HLA-II-associated peptides generally exhibited larger absolute MFE values, suggesting increased predicted transcript stability following optimisation, whereas HLA-I peptides showed broader dispersion across the CAI–MFE space. These results indicate that candidate neoantigens differ considerably in their manufacturability characteristics even prior to biological prioritisation. To integrate peptide-level optimisation into systems-level analyses, peptide optimisation metrics were aggregated into gene-level scores weighted by CAI × |MFE| and used as input for the Pi analysis (**Figure 3a,c**). Given the emerging importance of coordinated MHC-I and MHC-II neoantigen landscapes in shaping anti-tumour immunity and immunotherapy response^24^, gene-level prioritisation incorporated optimisation evidence from both HLA classes to favour candidates with the potential to simultaneously engage CD8□ and CD4□ T-cell responses. Several genes exhibited strong optimisation metrics across both HLA classes, including *BCR, BRAF, DDB1, RNPEP*, and *EIF4A2*.

**Figure 3.**
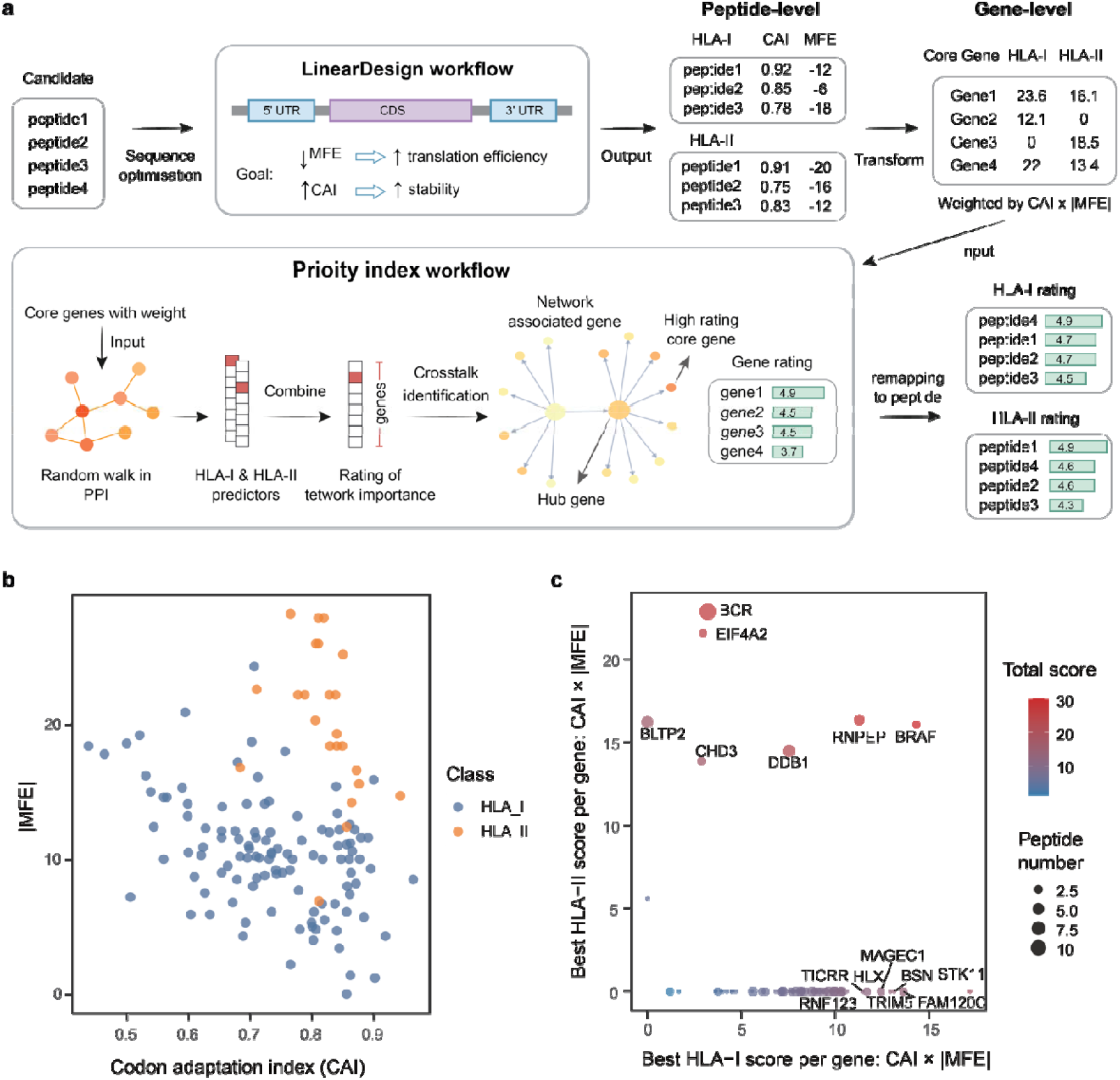
Integrated framework for mRNA sequence optimisation and gene-level prioritisation. (a) The schematic details the integrated PimRNA framework. First, mRNA sequence optimisation is applied to candidate neoantigen peptides using LinearDesign to maximise the codon adaptation index and minimise folding free energy. The resulting peptide-level metrics are then aggregated into gene-level scores weighted by the product of these two parameters and propagated through a protein-protein interaction network using a random walk with restart. Integrated HLA-I and HLA-II predictors are used to infer network importance and identify pathway crosstalk, generating prioritised gene rankings which are subsequently mapped back to the peptide level. (b,c) The peptide-level optimisation landscape is presented with each candidate peptide coloured by HLA class, followed by a gene-level aggregation where each point represents a gene, with point size indicating the number of associated peptides and colour denoting the total optimisation score across HLA classes; selected high-scoring genes are annotated.

### PimRNA prioritisation identifies network-central neoantigen-associated genes

Genes corresponding to candidate neoantigen peptides identified by NeoDisc were defined as core genes and subsequently used as seeds for random walk with restart analysis within a protein–protein interaction network, generating genome-wide HLA-I and HLA-II predictors. These predictors were then integrated into a unified priority rating to identify genes showing concordant evidence from neoantigen presentation and network-level functional connectivity. The resulting prioritisation network revealed several highly connected genes occupying central positions (**Figure 4**), including *BRAF, BCR, DDB1, EIF4A2, CHD3*, and *STK11*. Multiple prioritised genes have established roles in melanoma biology and cancer-associated signalling. For example, *STK11* functions as a tumour suppressor regulating AMPK–mTOR signalling and metabolic adaptation^24^, whereas *EIF4A2* regulates cap-dependent translation initiation, a process frequently dysregulated in cancer^25^. Notably, the *EIF4A2*-derived peptide was originally ranked 101st by NeoDisc but was reprioritised to 6th place in the PimRNA HLA-I ranking, illustrating the ability of network-informed prioritisation to rescue biologically relevant candidates overlooked by peptide-centric approaches. Collectively, these observations suggest that PimRNA prioritisation enriched for genes involved in functionally important tumour-associated regulatory processes.

**Figure 4.**
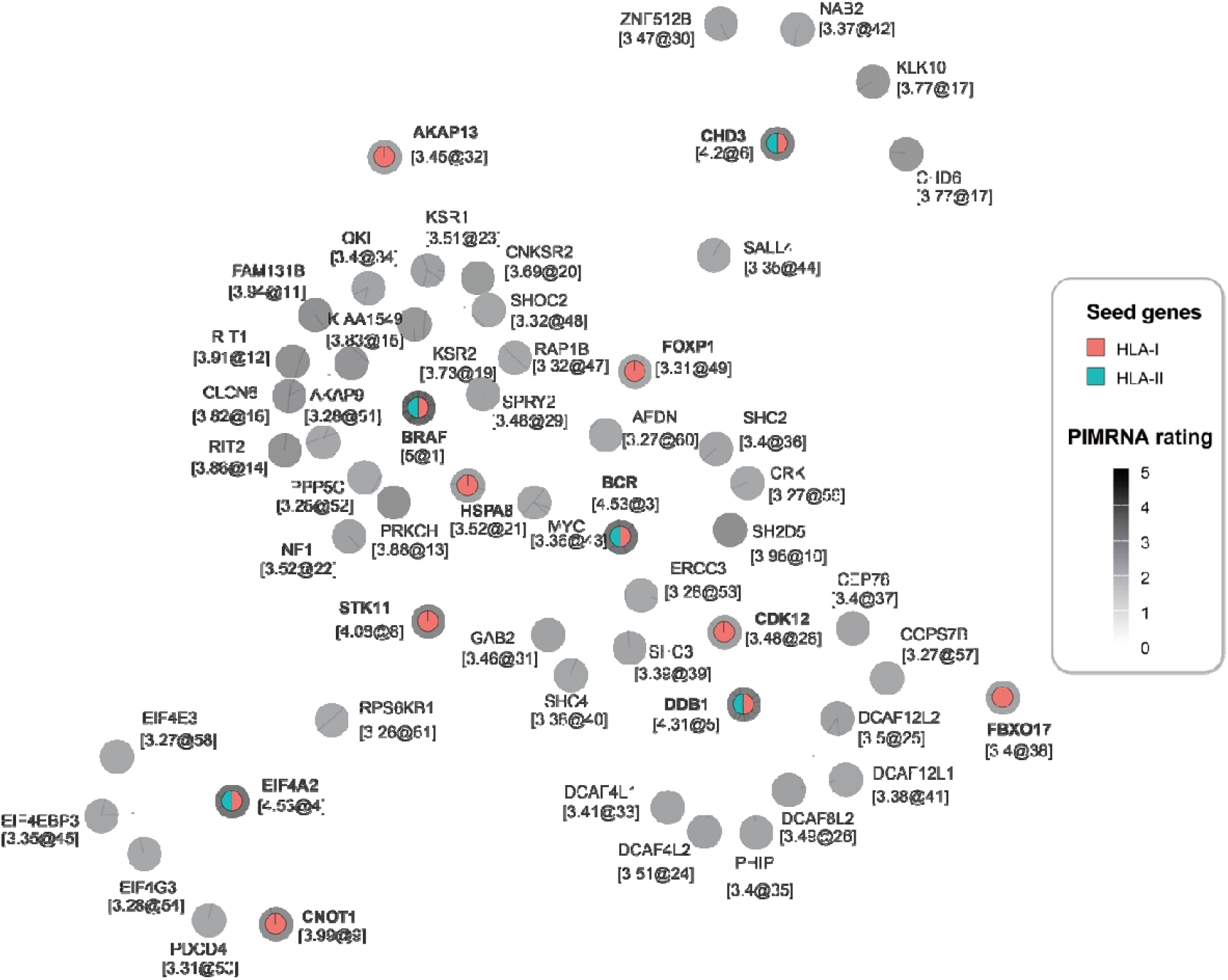
PimRNA prioritised gene interaction network of candidate neoantigen targets. The network illustrates functional associations between genes. Each node is labelled with its gene symbol, followed by its rating and rank in the format gene [rating@rank]. Node greyscale shading reflects the PimRNA prioritisation rating, with darker nodes indicating higher network importance. Seed genes are additionally denoted by coloured sectors, where red indicates HLA-I and cyan indicates HLA-II neoantigen support.

### Crosstalk analysis identifies signalling modules associated with melanoma progression

Tumour signalling pathways rarely operate in isolation, and therapeutically relevant neoantigens may preferentially arise from genes positioned at the intersection of multiple oncogenic processes. We therefore performed pathway crosstalk analysis using highly rated PimRNA-associated genes to identify subnetworks (**Figure 5a**). The resulting crosstalk network revealed densely interconnected signalling modules centred around hub genes including *BRAF* and *DDB1*, suggesting that these genes coordinate communication between multiple functional pathways. Pathway-level analysis demonstrated that crosstalk-associated genes were enriched in several canonical oncogenic signalling pathways, including MAPK, PI3K–Akt, ErbB, Ras, Rap1, mTOR, and cAMP signalling (**Figure 5b**). Many of these pathways are strongly implicated in melanoma pathogenesis and therapeutic resistance. In particular, aberrant MAPK signalling driven by oncogenic *BRAF* mutations represents a defining molecular feature of melanoma and constitutes a major therapeutic target in current clinical practice^25^. Similarly, PI3K-Akt and mTOR signalling pathways contribute to tumour survival, metabolic adaptation, immune evasion, and resistance to MAPK-targeted therapies^26^.

**Figure 5.**
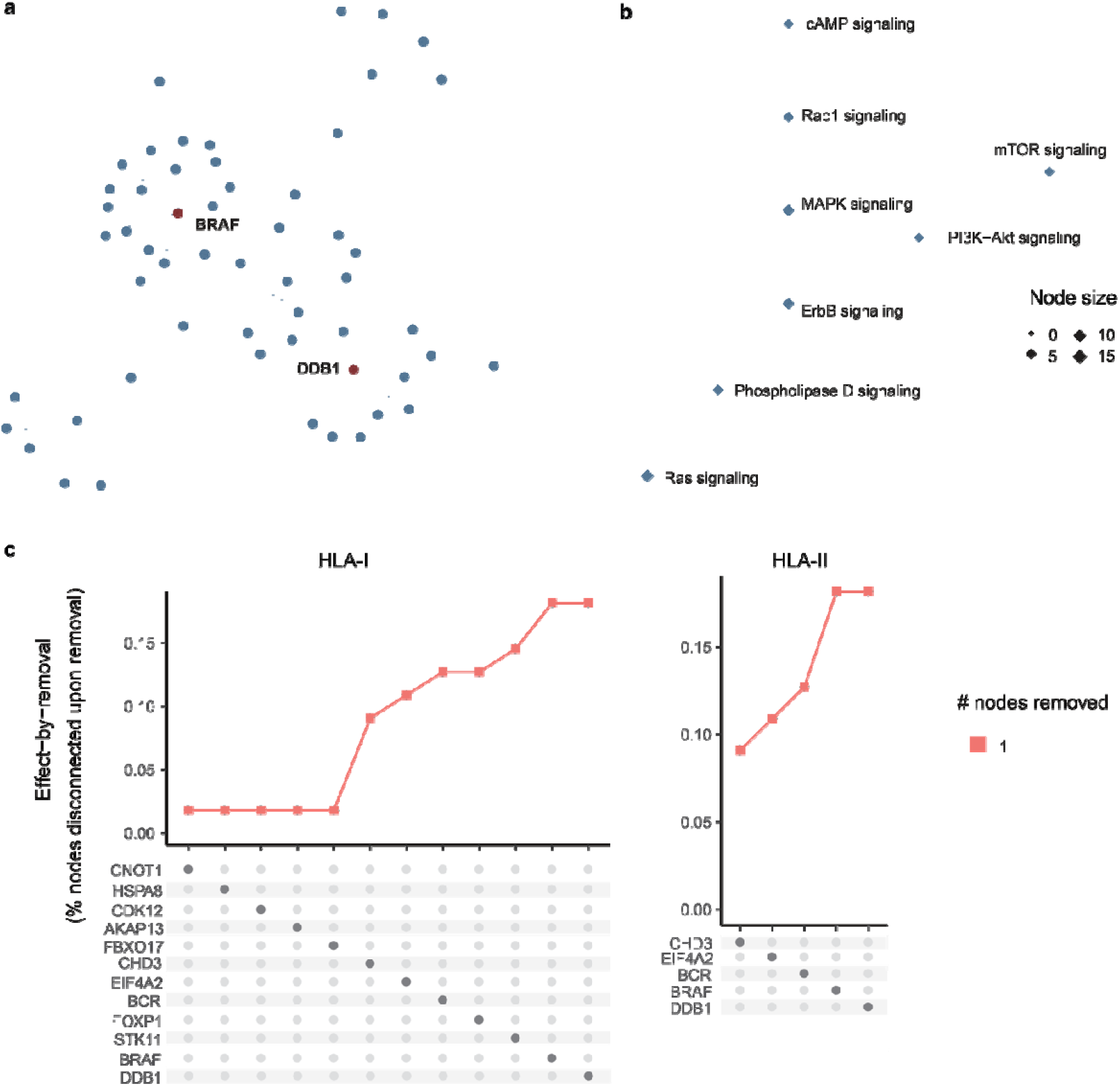
Network crosstalk and perturbation analysis of PimRNA prioritised neoantigen-associated genes. Panel (a) shows the crosstalk among high-priority genes within KEGG pathways, where hub genes form densely connected subnetworks linked to multiple functional modules, indicating their central role in coordinating tumour signalling. Panel (b) displays the functional pathway associations of these genes, with node size representing the number of associated genes contributing to each pathway. Panel (c) presents a network perturbation analysis evaluating the impact of sequential gene removal, where the upper panels show the proportion of disconnected nodes and the lower matrices detail the specific genes removed at each step. The loss of network integrity upon removal of these prioritised genes supports their central role as neoantigen-associated candidates.

### Perturbation analysis reveals structural dependence on prioritised hub genes

To evaluate the structural importance of prioritised genes within the crosstalk network, a perturbation-based effect-by-removal analysis was performed (**Figure 5c**). In the HLA-I network, removal of genes such as *DDB1, BRAF, EIF4A2, CHD3*, and *FBXO17* produced marked increases in the proportion of disconnected nodes, suggesting that these genes function as major organisational hubs within the interaction network. Similarly, the HLA-II network exhibited pronounced sensitivity to removal of central genes including *DDB1, BRAF*, and *EIF4A2*. These findings support the hypothesis that targeting neoantigens derived from network-central genes may exert broader functional consequences than targeting biologically peripheral antigens. Together, these findings suggest that integrating immunogenicity with network-informed functional prioritisation may improve the identification of biologically meaningful neoantigen targets for mRNA cancer vaccine development.

## Discussion

Our PimRNA framework not only refines candidate neoantigen selection but also enables gene-level visualisation of functional relationships among predicted peptides, facilitating the identification of tumour-associated subnetworks with potentially greater therapeutic relevance. Importantly, several hub genes identified within network crosstalk were not always associated with top-ranked neoantigenic peptides, yet may still represent important oncogenic drivers. In the context of multivalent mRNA vaccine design, neoantigens derived from such genes could potentially be co-formulated alongside highly immunogenic but biologically less connected neoantigen candidates to broaden therapeutic coverage and may reduce antigen-loss-mediated immune escape, as loss of these targets may impose substantial fitness costs on tumour cells.

Several limitations should be acknowledged. The current framework relies primarily on computational predictions of neoantigen presentation, network connectivity, and mRNA sequence optimisation, without direct experimental validation of translational efficiency or immunogenicity. Furthermore, the weighting strategy based on CAI and MFE represents a simplified approximation of mRNA manufacturability and may not fully capture the complexity of in vivo mRNA stability or innate immune activation. In addition, the present study was evaluated in a limited number of tumour samples. Future studies incorporating larger patient cohorts, single-cell tumour ecosystems, spatial transcriptomics, and experimental validation will be important for establishing the clinical utility of systems-informed mRNA neoantigen prioritisation.

Looking ahead, the peptidein landscape offers an unexplored dimension for PimRNA. By incorporating peptidein-encoding sequences into mRNA vaccine constructs, it becomes possible to target an entirely new class of tumour antigens, potentially enhancing vaccine efficacy in settings where somatic mutations alone provide insufficient immunogenic targets. This approach would be especially valuable in hard-to-treat, low-mutation-burden cancers, where peptideins could supply the antigenic diversity required to elicit robust, durable T-cell responses. The integration of peptidein-derived neoantigens with PimRNA’s network-based prioritisation framework therefore represents a logical next step toward comprehensive, systems-informed mRNA vaccine design.

Beyond its application as a standalone prioritisation framework, PimRNA may also provide a computational foundation for future agentic systems in precision mRNA therapeutics. Recent demonstrations of language model-based research agents—such as Virtual Lab, which autonomously designed novel nanobodies^27^, and the “AI co-scientist” that generated experimentally validated hypotheses^28^—highlight the potential for intelligent systems to orchestrate complex biomedical workflows. In this context, we have previously conceptualised the PimRNAgent framework^29^, a multi-agent system wherein specialised agents for task analysis, planning, execution, and critique collaborate to design personalised mRNA vaccines. PimRNA could serve as a core analytical module within such a system by integrating neoantigen discovery, mRNA optimisation, and systems-level network analysis into a unified prioritisation framework. Overall, by integrating network biology into the target selection process, PimRNA provides a generalisable strategy for systems-informed prioritisation of personalised mRNA therapeutics.

## Data Availability

All data produced in the present study are available upon reasonable request to the authors

